# Recording of patient-reported outcome measures for anxiety disorders and depression in English primary care: An analysis of public data using OpenCodeCounts

**DOI:** 10.64898/2026.11.26.26368944

**Authors:** Lola Ojedele, Kunle Oreagba, Yamina Boukari, Helen J. Curtis, Rob Saunders, Tony Kendrick, David M. Clark, Graham R. Thew, Arina A. Tamborska, Rose Higgins, Brian MacKenna, Ben Goldacre, Sebastian Bacon, Milan Wiedemann

## Abstract

**Background:** Patient-reported outcome measures (PROMs) are widely used to assess symptom severity, guide treatment, and inform service improvement for anxiety disorders and depression. However, PROMs use is not systematic in primary care, where most individuals first present with these conditions.

**Aim:** To describe PROMs recording from August 2011 to July 2026 for anxiety disorders and depression in primary care electronic health records in England.

**Design and setting:** Retrospective descriptive analysis of publicly available national SNOMED CT code usage data from most general practices in England through OpenCodeCounts.

**Method:** We developed PROMs activity and declined-assessment codelists for anxiety disorders and depression, described code usage, and plotted annual trends.

**Results:** Over the past 5 years, GAD-7 and PHQ-9 were the most frequently recorded PROMs for anxiety disorders and depression, respectively. Recording of brief screening questionnaires, including PHQ-2 and GAD-2, also increased alongside their longer versions since 2019. Recording shifted from PROMs procedure codes, towards codes that record PROMs scores. Depression-related PROMs codes were recorded around 4 times as often as anxiety disorder-related codes (49,611,450 vs 13,783,300) across the study period, decreasing to 1.5 times in July 2026 (4,343,895 vs 2,873,410). PHQ-9 score usage dropped by half after financial incentives ended in 2013, recovering completely by 2026.

**Conclusion:** Publicly available aggregate data can describe PROMs recording trends and inform feasibility of research on anxiety disorders and depression in English general practice.

**How this fits in:** The National Institute for Health and Care Excellence (NICE) and the British Medical Association (BMA) recommend the use of patient reported outcome measures (PROMs) for assessing symptom severity in anxiety disorders and depression. How often and in what form these measures are recorded in general practice is unclear. We used OpenCodeCounts, an openly accessible online tool and R package that shows how often clinical codes are used in England. Over the past 5 years, recording shifted from codes that only indicate whether a questionnaire was given, towards codes that record the measure scores, specifically PHQ-9 and GAD-7.

Additionally, recording of PHQ-9 declined after financial incentives ended in 2013, before increasing again by the end of the study period in 2026. These findings provide national context on PROMs recording in primary care. They also show how publicly available data can inform future patient-level studies.

## Introduction

Anxiety disorders and depression are the most common mental health conditions in England. The nationally representative Adult Psychiatric Morbidity Survey found that the prevalence among adults aged 16-64 increased between 1993 and 2023/24 from 15.5% to 20.2% [1]. Recorded anxiety disorder diagnoses and symptoms in primary care have similarly increased [2,3]. Primary care is the main point of contact for people with anxiety disorders and depression in England [1]. Psychological (talking) therapies are recommended for these conditions, with most such therapy in England provided by NHS Talking Therapies (NHS TT) for anxiety and depression, which treated over 674,000 adults in 2025-26 [4,5]. In both settings, structured assessment of symptom severity guides clinical decisions, including identifying appropriate treatment and monitoring progress [6,7].

Patient-reported outcome measures (PROMs) are self-completed questionnaires that assess symptom severity and daily functioning in physical and mental health conditions. They are used across primary, secondary, and community-based care [8]. The National Institute for Health and Care Excellence (NICE) and the British Medical Association (BMA) recommend validated measures to assess severity and guide treatment decisions [6,7,9]. For depression, the Patient Health Questionnaire-9 [PHQ-9, 10], the Beck Depression Inventory [BDI-II, 11], and the depression subscale of the Hospital Anxiety and Depression Scale [HADS-D, 12] are recommended for symptom severity assessment. Brief screening questionnaires for depression include the Whooley screening [13] and the 2-item version of the PHQ [PHQ-2, 14]. Both these measures ask the same questions, but differ on response options (yes/no vs Likert scale). For anxiety disorders, the Generalised Anxiety Disorder 7-item scale [GAD-7, 15] is widely used for symptom severity, with the 2-item version (GAD-2) used for screening [6]. How these recommendations translate into routine practice is less clear.

Systematic use of PROMs at initial assessment and follow-up can improve patient outcomes, particularly in psychological therapy settings [4,16–19]. In primary care, the Quality and Outcomes Framework (QOF) financially incentivised general practitioners to record structured severity assessments for depression at diagnosis from 2006 and at follow-up from 2009, until these indicators were retired in 2013 [17,20]. However, evidence on the impact of PROMs use on patient outcomes in primary care remains limited. Further research on patient outcomes first requires identifying how PROMs are recorded in primary care electronic health records (EHR), including which measures are used and how often [21].

We set out to describe PROMs recording and use for anxiety disorders and depression in English general practice using publicly available aggregated data from NHS England [22]. This study aims (1) to develop open codelists for PROMs based on national guidance, (2) to describe how often and in what form these measures are recorded over time, and (3) to show how publicly available aggregate data can inform future patient-level studies.

## Methods

### Design and setting

This was a retrospective descriptive analysis of PROMs-related coding in primary care EHRs. We analysed national SNOMED CT code usage across all general practices in England that contributed to the public dataset from August 2011 to July 2026. The dataset included 6,565 general practices with 63,665,066 registered patients in 2025-26.

### Data sources

We used publicly available [SNOMED CT Code Usage in Primary Care](https://digital.nhs.uk/data-and-information/publications/statistical/mi-snomed-code-usage-in-primary-care) data from NHS England, accessed as dataset version 0.2.0 through the *opencodecounts* R package [Version 0.10.0, 22]. SNOMED CT is a standardised clinical terminology that gives each concept a code, a description and a semantic tag for the type of information. For example, “observable entity” codes record a measurable result (e.g., questionnaire score), and “procedure” codes record an activity (e.g., undertaken assessment). The usage count reflects the number of times each listed SNOMED CT code was added to a patient’s primary care record in England during the annual reporting period, with counts rounded to the nearest 10 and counts of five or below displayed as 5. Each yearly data point is labelled by its end date (e.g., “July 2024” covers 1 August 2023 to 31 July 2024).

### Codelist development

Codelists were created following guidance on codelist development [21]. We identified search terms and specific SNOMED CT codes from NICE guidelines [6,7,23,24], the NHS TT manual [25], and the [NHS Primary Care Domain Reference Set](https://digital.nhs.uk/data-and-information/data-collections-and-data-sets/data-collections/quality-and-outcomes-framework-qof/quality-and-outcome-framework-qof-business-rules/primary-care-domain-reference-set-portal) (Supplementary Table S1). We also consulted a general practitioner to identify additional clinically relevant search terms. For each condition, two codelists were developed: one capturing PROMs activity codes indicating completed assessments and recorded scores, and one capturing declined assessments. All four codelists developed in this study are published on OpenCodelists and available for reuse (Supplementary Table S2).

#### PROMs for anxiety disorders

The activity codelist included 76 SNOMED CT codes, including the GAD-7 [15], GAD-2 [26], the anxiety subscale of the HADS [HADS-A, 12], and anxiety disorder-specific measures, such as the PTSD Checklist for DSM-5 [PCL-5, 27] and Social Phobia Inventory [SPIN, 28]. The declined assessments codelist included 3 codes indicating that an assessment was offered but not completed.

#### PROMs for depression

The activity codelist included 56 SNOMED CT codes, including the PHQ-9 [10], PHQ-2 [14], BDI-II [11], and HADS-D [12], as well as broader depression screening concepts. The declined assessments codelist included 5 codes.

### Data analyses

We combined the usage of nine pairs of equivalent codes identified through [SNOMED CT history records](https://isd.digital.nhs.uk/trud/users/guest/filters/0/categories/8/items/276/releases) or manual review. We identified the 10 most frequently recorded codes for the activity and declined codelists for anxiety disorders and depression PROMs. We plotted the annual activity broken down by semantic tag including “observable entity”, “procedure”, and all other tags. Annual usage counts for the 3 most frequently recorded activity codes were also plotted. All analysis code and clinical codelists are openly available at https://github.com/bennettoxford/proms-primary-care-public-data.

## Results

### Total PROMs code usage across the study period

We identified 13,783,300 anxiety disorder-related and 49,611,450 depression-related PROMs recording instances between August 2011 and July 2026. For both condition groups, almost all recording instances were captured by the 10 most frequently used codes, which represented 99.4% and 98.8% of recording instances for anxiety disorders and depression respectively (Table 1).

**Table 1.** Top ten recorded SNOMED CT PROMs codes for anxiety disorders and depression in England from August 2011 to July 2026.

| **Description** | **Semantic tag** | **Usage** | **%** |
|---|---|---|---|
| **PROMs codes for anxiety disorders** |  |  |  |
| GAD-7 score | Observable entity | 9,196,590 | 66.7 |
| GAD-2 score | Observable entity | 2,744,860 | 19.9 |
| HADS anxiety score | Observable entity | 800,120 | 5.8 |
| Anxiety screening | Procedure | 486,195 | 3.5 |
| GAD-2 | Assessment scale | 125,070 | 0.9 |
| PHQ-4 score | Observable entity | 114,225 | 0.8 |
| Anxiety screening using questions | Procedure | 87,460 | 0.6 |
| HADS | Assessment scale | 61,120 | 0.4 |
| Anxiety screening declined | Situation | 46,510 | 0.3 |
| Assessment using GAD-7 declined | Situation | 38,100 | 0.3 |
| **PROMs codes for depression** |  |  |  |
| Depression screening using questions | Procedure | 23,096,200 | 46.6 |
| PHQ-9 score | Observable entity | 20,384,150 | 41.1 |
| PHQ-2 score | Observable entity | 2,946,700 | 5.9 |
| HADS depression score | Observable entity | 867,030 | 1.7 |
| Depression screening | Procedure | 847,890 | 1.7 |
| Depression screening declined | Situation | 200,900 | 0.4 |
| EPDS score | Observable entity | 198,280 | 0.4 |
| EQ-5D-5L anxiety depression score | Observable entity | 175,055 | 0.4 |
| Whooley depression screening | Procedure | 164,910 | 0.3 |
| PHQ-4 score | Observable entity | 114,225 | 0.2 |

*Notes*. GAD = Generalised Anxiety Disorder; HADS = Hospital Anxiety and Depression Scale; PHQ = Patient Health Questionnaire; EPDS = Edinburgh Postnatal Depression Scale; EQ-5D-5L = EuroQol five dimension five level. % = Percentage usage for each PROMs code is calculated separately for anxiety disorders and depression, using all recorded events for that condition as the denominator.

In July 2026, anxiety disorder PROMs recording was predominantly represented by observable entity concepts (94.8%), such as “*GAD-7 score*” and “*GAD-2 score*”. Depression PROMs recording was also mainly represented by observable entity concepts (78.0%), with fewer procedure concepts (21.2%), such as “*Depression screening using questions*”. Situation, finding, and assessment scale semantic tag concepts accounted for 0.5% of anxiety disorder and 0.8% of depression recorded events.

### Annual trends in overall PROMs recording

Annual trends showed higher depression-related PROMs coding than anxiety disorder-related coding throughout the study period. However, depression-related PROMs coding declined from 6,150,730 recorded events in July 2012 to 2,856,095 by July 2014, increasing slowly to 4,343,895 in July 2026. In contrast, anxiety disorder PROMs recording remained lower but increased over time, reaching 2,873,410 recorded events by July 2026 (Figure 1, Panel A).

**Figure 1.**
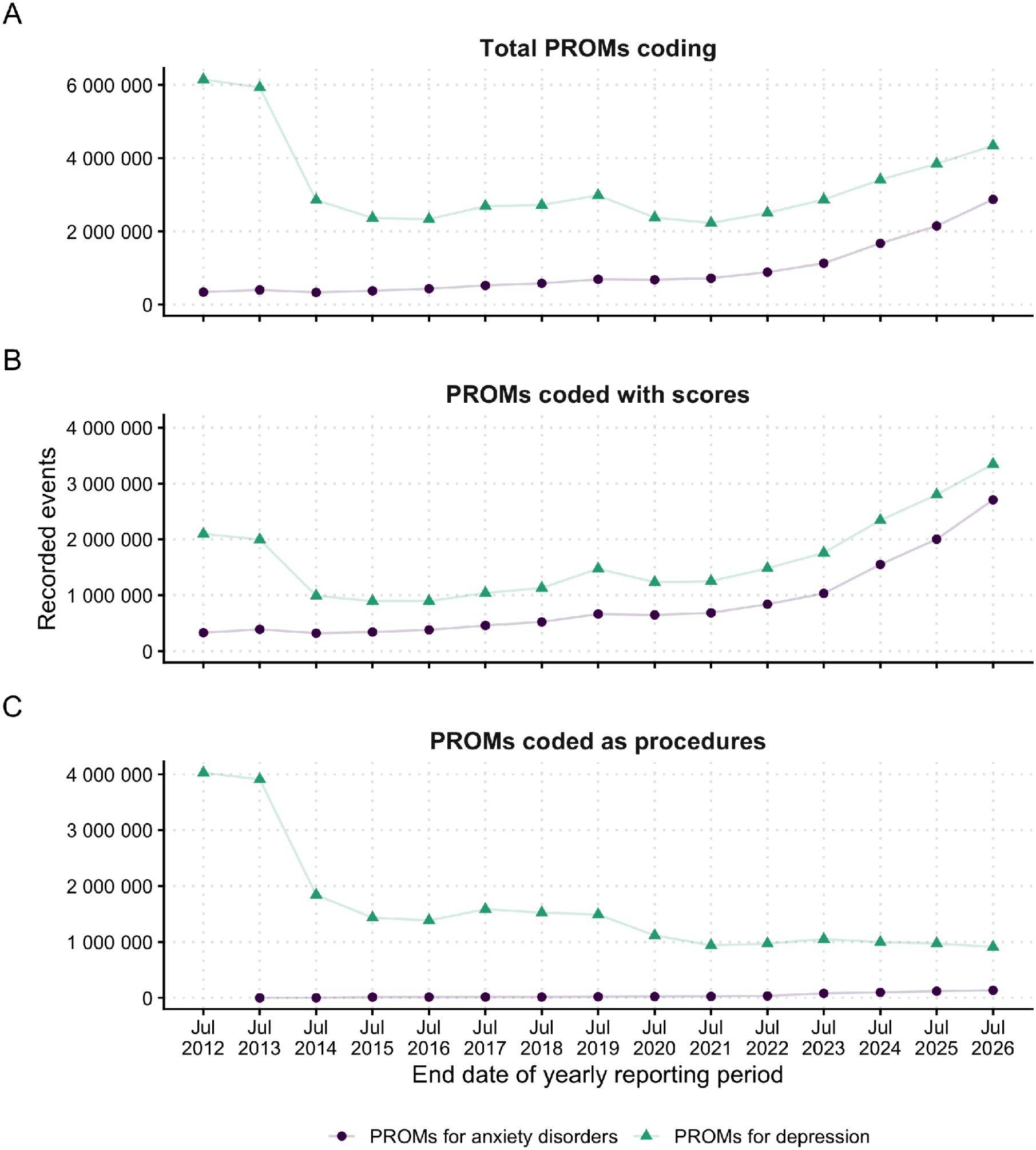
Yearly recorded events for PROMs codes for anxiety disorders and depression in English general practice from August 2011 to July 2026. (A) Total PROMs coding, including all activity and declined codes; (B) score codes, which are observable entity concepts representing recorded PROM scores; and (C) procedure codes, representing PROM administration. No procedure code events were recorded for anxiety disorders before 2013. Panels B and C share the same y-axis range, which differs from Panel A.

PROMs for depression were initially most often recorded as procedures, but this declined substantially from 4,028,320 in July 2012 to 912,090 in July 2026 (Figure 1, Panel C). Depression-related PROMs with scores (observable entity) declined from 2,098,905 in July 2012 to 897,490 in July 2016, before increasing again to 3,348,960 by July 2026 (Figure 1, Panel B). Anxiety disorder PROMs were rarely recorded as procedures, with no more than 134,910 recorded events in any year, while codes with scores increased from 330,475 in July 2012 to 2,708,760 in July 2026 (Figure 1, Panel B-C). Codes indicating that patients declined recording of PROMs increased over the study period but remained low, reaching at most 47,785 recorded events in any year for depression and 16,680 for anxiety disorders (Supplementary Figure S1). This represented 1 in 112 recorded PROMs codes by July 2026.

### Annual trends in the most used individual PROMs codes

Figure 2 shows yearly trends in recorded events for the 3 most frequently recorded PROMs SNOMED CT codes.

**Figure 2.**
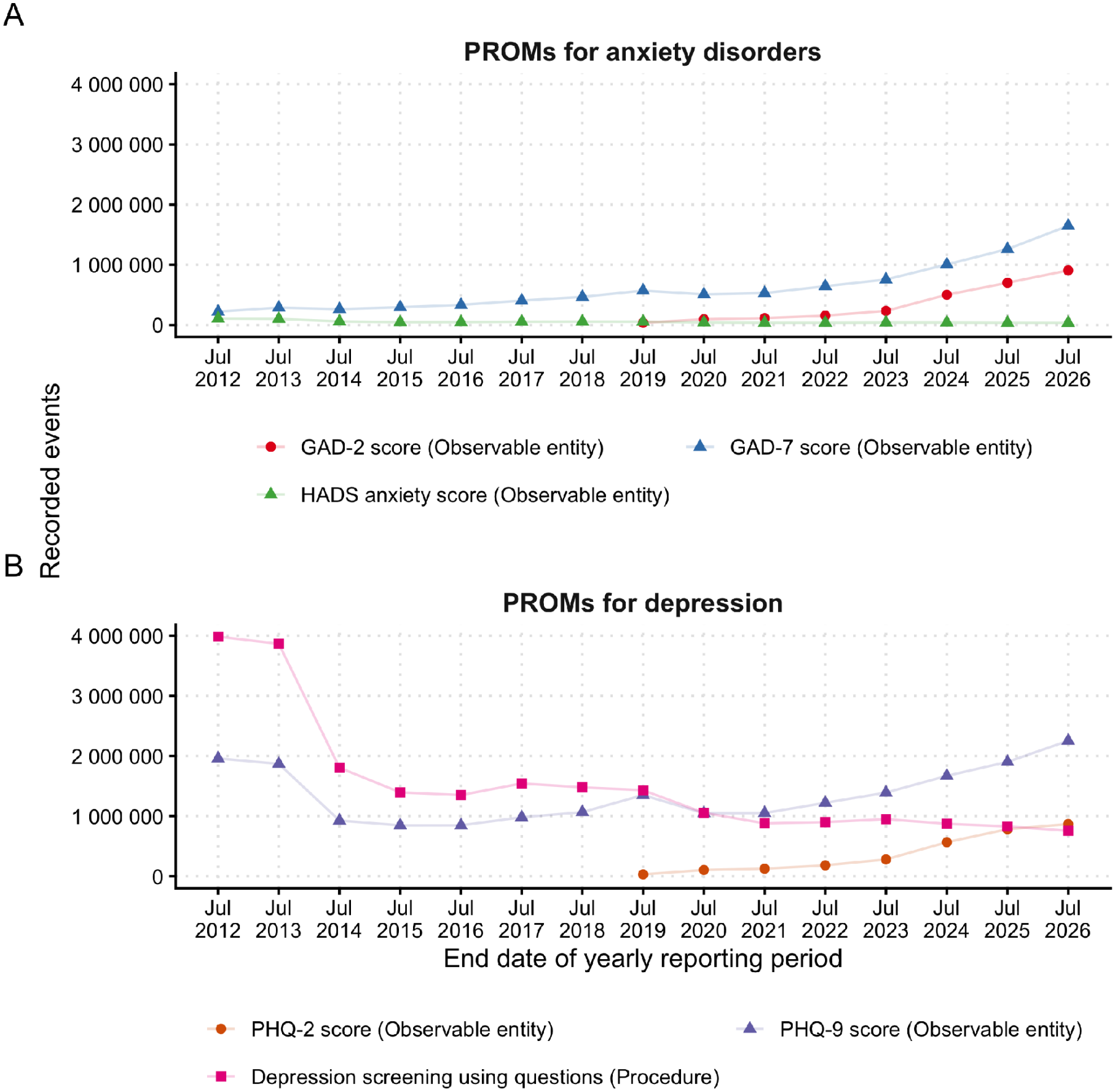
Yearly recorded events for the 3 most frequently recorded PROMs activity codes for anxiety disorders and depression in English general practice from August 2011 to July 2026. Circles show brief screening questionnaires (GAD-2 and PHQ-2). Triangles show long-form questionnaires (PHQ-9, GAD-7, and the HADS anxiety subscale). Squares show procedure codes, which do not identify the questionnaire used.

Among the PROMs codes for anxiety disorders, “*GAD-7 score*” was the most frequently recorded code throughout the study period and increased overall from July 2012 to July 2026 (Figure 2, Panel A). “*GAD-2 score*” recording appeared from July 2019 and increased thereafter. Both codes showed their most rapid increases from 2023. The remaining code (“*HADS anxiety score*”) was recorded less often, with no more than 107,810 recorded events per year.

For the two most frequently used depression-related PROMs codes, recording declined between July 2012 and July 2014. However “*Depression screening using questions*” continued to decrease to 756,670 by July 2026, whereas “*PHQ-9 score*” increased again to 2,252,480, above its level in July 2012, the year before the QOF financial incentives for using PROMs were discontinued (Figure 2, Panel B). “*Depression screening using questions*” was overtaken for the first time by “*PHQ-9 score*” in July 2021 and by “*PHQ-2 score*” in July 2026. “*PHQ-2 score*” was first recorded in July 2019 and increased to 868,220 by July 2026.

Recording of brief screening questionnaires, including PHQ-2 and GAD-2, increased alongside recording of PHQ-9 and GAD-7. Brief screening questionnaires (PHQ-2, GAD-2, PHQ-4, and Whooley questions) accounted for 36.0% of anxiety disorder and 23.7% of depression activity coding in July 2026, up from 0.0% and 0.1% in July 2012.

## Discussion

### Summary

Over the past 5 years, GAD-7 and PHQ-9 were the most frequently recorded PROMs annually for anxiety disorders and depression respectively. While “*Depression screening using questions*” had the highest cumulative recording across the study period, “*PHQ-9 score*” was the most frequently recorded code annually from July 2021 onwards. Depression-related PROMs coding was consistently higher than anxiety disorder-related coding. Over the study period, recording shifted from broader procedure codes towards named PROMs score codes, most evident for depression.

### Comparison with existing literature

Consistent with previous research on depression severity assessments [20], most recording was captured by a small number of PROMs. PHQ-9 and GAD-7 were the most recorded named measures in recent years, consistent with NICE recommendations [6,7]. Their frequent recording may also reflect their role as standard outcome measures in NHS TT [25]. The high usage of generic depression screening codes is also consistent with reported variation in PROMs recording across general practice [29]. Although NICE recommends anxiety disorder-specific measures for conditions such as social anxiety disorder [23], these instruments were rarely recorded in general practice. NICE also recommends the 3-item mini-SPIN [30] to identify possible cases of social anxiety [23]. We could not identify a SNOMED CT code for the mini-SPIN, so its use cannot be recorded in a structured form. In contrast, the brief PHQ-2 and GAD-2 have their own codes, and their recording increased alongside their longer versions. However, these are primarily used for screening [6,14,26], so their increase may not reflect more frequent outcome monitoring.

Depression PROMs coding was consistently higher than anxiety disorder PROMs coding across the study period, despite anxiety disorders being at least as common in the community [1]. This potentially reflects the earlier financial incentives to use depression PROMs under QOF [17,20]. The early decline of depression coding coincided with the removal of these incentives in 2013 [17,20]. The frequent recording of PHQ-9 is consistent with patterns observed under QOF, where it accounted for most recorded depression severity assessments [20]. NICE guidance continued to recommend routine outcome monitoring [7], which may partly explain why PHQ-9 continued to be recorded. By 2026, PHQ-9 score recording had recovered to levels above those observed under QOF incentivisation. Possible explanations include psychological services placing greater value on symptom scores for referred patients, or changes in how symptom questionnaires are used in primary care. For example, recorded remote consultations in English general practice more than tripled during the COVID-19 pandemic [31]. Patient communication platforms enable patients to complete PROMs remotely, with scores written into the patient’s EHR, including PHQ-9 and GAD-7 scores [32]. For anxiety disorders, GAD-7 coding increased over the study period, while GAD-2 score recording increased from 2019. This parallels broader increases in recorded anxiety diagnoses and symptoms in UK primary care from around 2011 [2,3].

In 2016, a Cochrane review found insufficient evidence that routine PROMs monitoring improves outcomes in common mental health disorders, with only two of 17 studies conducted in primary care [16]. Since then, a cluster randomised trial in English and Welsh general practices found no significant effect of PHQ-9 monitoring on depression symptom outcomes, although patients’ quality of life improved [18].

Given the substantial growth in PROMs recording in general practice since 2016, national studies using patient-level data could first establish how PROMs are used for repeated monitoring and then examine their relationship with patient outcomes.

### Strengths and limitations

Strengths of this study include its national scale and the effective use of publicly available aggregate data without requiring patient level access. It demonstrates the combined use of OpenCodelists and OpenCodeCounts [22], which may support future studies.

The main limitation is that this study used aggregate counts of recorded code usage, rather than patient-level data. This means we cannot determine how often PROMs were recorded for individual patients. It is also difficult to disentangle changes in clinical coding from changes in practice, especially when recording is not financially incentivised. Some PROMs-related information may be recorded in free text clinical notes rather than structured codes, so the analysis may underestimate PROMs use. Before 2019, data was submitted in Read v2 or CTV3 format and mapped to SNOMED CT, which may affect comparability over time. We also cannot tell whether brief screening questionnaires were used alone or recorded alongside a full measure. Generic codes, such as “*Depression screening using questions*”, do not identify which specific PROM was administered.

### Implications for research and practice

The increase in “*PHQ-9 score*” and “*GAD-7 score*” recording means that general practice now increasingly records the same measures used as outcomes in NHS TT. Patient-level linkage of primary care and NHS TT records would allow symptom severity to be analysed across both settings. The codelists developed in this study identify PROMs recording in primary care records and can be used for these analyses.

NHS England also publishes aggregated PROMs outcomes from NHS TT, where the PHQ-9, GAD-7, and Work and Social Adjustment Scale [WSAS, 33] are collected as standard measures at every treatment session [25]. These measures are recorded for 98% of patients who received treatment with a score recorded at the start and end of treatment. The published outcomes are broken down by therapy type and main mental health problem [4]. In psychological therapy services there is good evidence that routine outcome monitoring improves patient outcomes [17,34]. The *nhstt* R package [35] provides access to this data in an analysis-ready format and could be used to describe how symptom severity at the start and end of treatment has changed over time and varies across services.

## Conclusion

Recording of PROMs in English general practice has shifted towards named score codes, particularly PHQ-9 and GAD-7. Publicly available aggregate data can be used to describe these trends and inform future patient-level research.

## Data Availability

All data produced are available online at https://github.com/bennettoxford/proms-primary-care-public-data.

## CRediT authorship contribution statement

**Conceptualisation**: LO, KO, MW, YB, HJC, BG, BMK, RS, DMC. **Methodology**: LO, KO, YB, HJC, GRT, RS. **Software**: LO, AAT, RH, SB, MW. **Validation**: TK, BMK,

GRT, DMC, YB, HJC, RS. **Formal analysis**: LO, KO, MW. **Investigation**: LO, KO, TK, MW. **Data Curation**: LO, KO, YB, AAT, RH, MW. **Writing - Original Draft**: LO, MW, HJC, RS. **Writing - Review & Editing**: All authors. **Visualization**: LO, KO, MW. **Supervision**: YB, MW, HJC, SB. **Project administration**: LO, MW. **Funding acquisition**: SB, MW, DMC, BG, BMK.

## Acknowledgements

We thank Health Data Research UK (HDR UK) for coordinating the Health Data Science Black Internship Programme, through which Lola Ojedele and Kunle Oreagba began this work at the Bennett Institute for Applied Data Science.

## Supplementary material

**Table S1:**
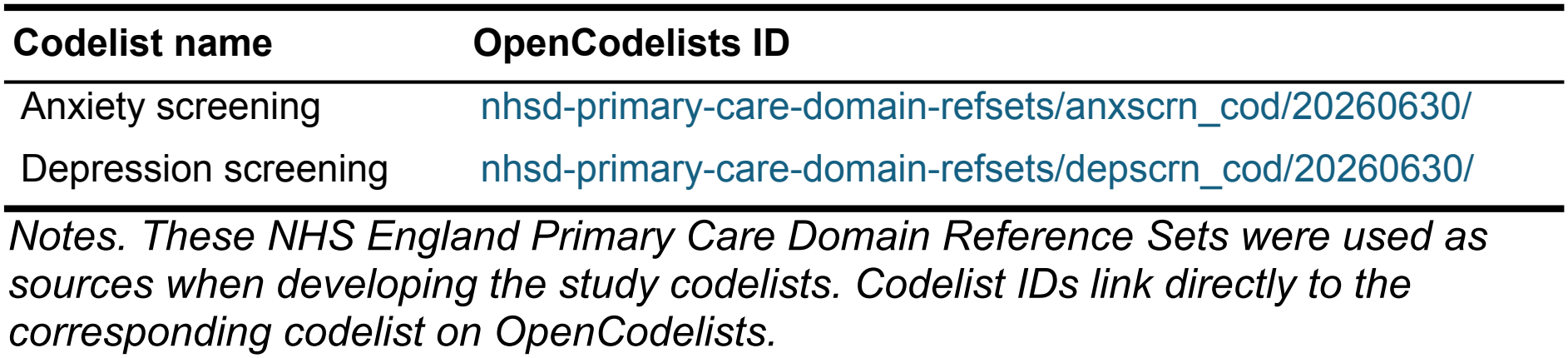
NHS England reference sets used to inform development of the anxiety disorder and depression PROMs codelists.

**Table S2:** PROMs activity and declined codelists for anxiety disorders and depression developed in this study.

| **Codelist name** | **OpenCodelists ID** |
|---|---|
| **Anxiety Disorders** |  |
| PROMs activity | user/Lola_O/proms-anx/1ed7683b/ |
| PROMs declined | user/Lola_O/proms-anx-declined/631bb866/ |
| **Depression** |  |
| PROMs activity | user/Lola_O/proms-dep/05c7f506/ |
| PROMs declined | user/Lola_O/proms-dep-declined/4a0c4531/ |

*Notes. Codelist IDs link directly to the corresponding codelist on OpenCodelists.The activity codelists share 3 codes, such as the Hospital Anxiety and Depression Scale, but these account for only 0.6% of anxiety disorder and 0.1% of depression recorded events.*

**Figure S1:**
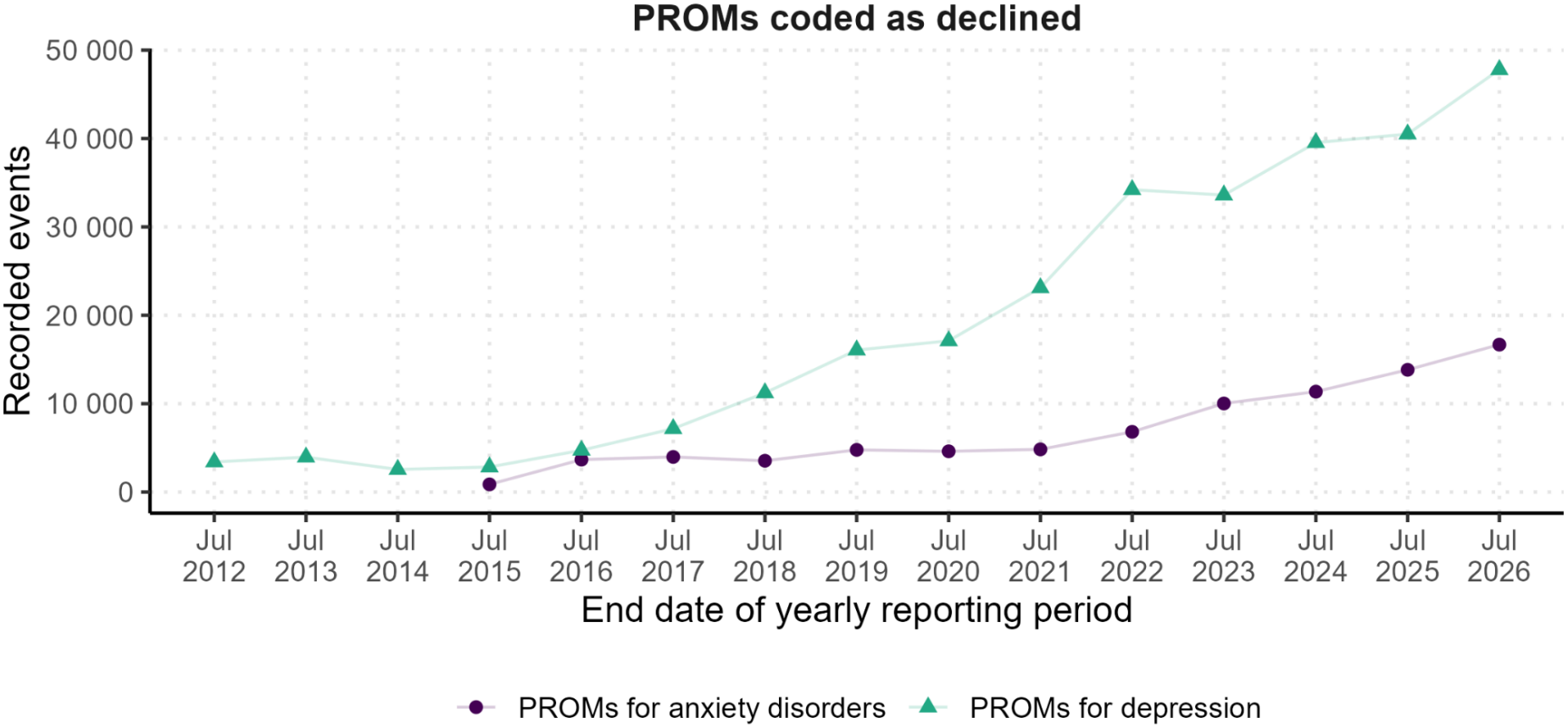
Yearly recorded events for declined PROMs codes for anxiety disorders and depression in English general practice from August 2011 to July 2026.

## References

[1] Morris S, Hill S, Brugha T, McManus S, editors. Adult Psychiatric Morbidity Survey-Survey of Mental Health and Wellbeing, England, 2023/4 2025. 10.13140/RG.2.2.24367.39840.

[2] Archer C, Turner K, Kessler D, Mars B, Wiles N. Trends in the recording of anxiety in UK primary care: A multi-method approach. Soc Psychiatry Psychiatr Epidemiol 2022;57:375–86. 10.1007/s00127-021-02131-8.

[3] Slee A, Nazareth I, Freemantle N, Horsfall L. Trends in generalised anxiety disorders and symptoms in primary care: UK population-based cohort study. Br J Psychiatry 2021;218:158–64. 10.1192/bjp.2020.159.

[4] Clark DM, Canvin L, Green J, Layard R, Pilling S, Janecka M. Transparency about the outcomes of mental health services (IAPT approach): An analysis of public data. The Lancet 2018;391:679–86. 10.1016/S0140-6736(17)32133-5.

[5] NHS England. NHS Talking Therapies, for anxiety and depression, Annual reports, 2025-26 2026. https://digital.nhs.uk/data-and-information/publications/statistical/nhs-talking-therapies-for-anxiety-and-depression-annual-reports/2025-26 (accessed November 5, 2026).

[6] National Institute for Health and Care Excellence. Generalised anxiety disorder and panic disorder in adults: management. NICE Clinical Guideline CG113. NICE; 2020.

[7] National Institute for Health and Care Excellence. Depression in adults: Treatment and management. NICE Clinical Guideline NG222. NICE; 2022.

[8] Kendir C, Tran S, Van Den Berg M, De Bienassis K. PROMoting quality of care through patient reported outcome measures (PROMs): Systematic collection of PROMs for quality improvement and assurance in 38 countries. 183rd ed. 2025. 10.1787/c17bb968-en.

[9] British Medical Association. Revisions to the GMS contract, 2006/07. Delivering Investment in General Practice. 2006.

[10] Kroenke K, Spitzer RL, Williams JBW. The PHQ-9: Validity of a brief depression severity measure. J Gen Intern Med 2001;16:606–13. 10.1046/j.1525-1497.2001.016009606.x.

[11] Beck AT, Steer RA, Brown GK. Beck Depression Inventory–II Manual. 2nd ed. Boston, MA: Hartcourt Brace; 1996.

[12] Zigmond AS, Snaith RP. The Hospital Anxiety and Depression Scale. Acta Psychiatr Scand 1983;67:361–70. 10.1111/j.1600-0447.1983.tb09716.x.

[13] Whooley MA, Avins AL, Miranda J, Browner WS. Case-finding instruments for depression. J Gen Intern Med 1997;12:439–45. 10.1046/j.1525-1497.1997.00076.x.

[14] Kroenke K, Spitzer RL, Williams JBW. The Patient Health Questionnaire-2: Validity of a Two-Item Depression Screener. Med Care 2003;41:1284–92. 10.1097/01.MLR.0000093487.78664.3C.

[15] Spitzer RL, Kroenke K, Williams JBW, Löwe B. A brief measure for assessing Generalized Anxiety Disorder: The GAD-7. Arch Intern Med 2006;166:1092. 10.1001/archinte.166.10.1092.

[16] Kendrick T, El-Gohary M, Stuart B, Gilbody S, Churchill R, Aiken L, et al. Routine use of patient reported outcome measures (PROMs) for improving treatment of common mental health disorders in adults 2016. 10.1002/14651858.CD011119.pub2.

[17] Kendrick T, Maund E. Do PROMS improve outcomes in patients with depression in primary care? BMJ 2020;370:m3313. 10.1136/bmj.m3313.

[18] Kendrick T, Dowrick C, Lewis G, Moore M, Leydon GM, Geraghty AW, et al. Depression follow-up monitoring with the PHQ-9: An open cluster-randomised controlled trial. Br J Gen Pract 2024;74:e456–65. 10.3399/BJGP.2023.0539.

[19] Gelkopf M, Mazor Y, Roe D. A systematic review of patient-reported outcome measurement (PROM) and provider assessment in mental health: Goals, implementation, setting, measurement characteristics and barriers. Int J Qual Health Care 2022;34:ii13–27. 10.1093/intqhc/mzz133.

[20] Kendrick T, Dowrick C, McBride A, Howe A, Clarke P, Maisey S, et al. Management of depression in UK general practice in relation to scores on depression severity questionnaires: Analysis of medical record data. BMJ 2009;338:b750. 10.1136/bmj.b750.

[21] Matthewman J, Andresen K, Suffel A, Lin L-Y, Schultze A, Tazare J, et al. Checklist and guidance on creating codelists for electronic health records research. NIHR Open Res 2024;4:20. 10.3310/nihropenres.13550.2.

[22] Tamborska AA, Higgins R, Boukari Y, Kingsley V, Ojedele L, Oreagba K, et al. OpenCodeCounts: An open-access, interactive online tool and R package for analysing clinical code usage in England 2025:2025.10.14.25338005. 10.1101/2025.10.14.25338005.

[23] National Institute for Health and Care Excellence. Social anxiety disorder: Recognition, assessment and treatment. 2013.

[24] National Institute for Health and Care Excellence. Post-traumatic stress disorder. NICE Clinical Guideline NG116. 2018.

[25] National Collaborating Centre for Mental Health. NHS Talking Therapies for anxiety and depression Manual. 2024.

[26] Kroenke K, Spitzer RL, Williams JBW, Monahan PO, Löwe B. Anxiety Disorders in Primary Care: Prevalence, Impairment, Comorbidity, and Detection. Ann Intern Med 2007;146:317–25. 10.7326/0003-4819-146-5-200703060-00004.

[27] Weathers FW, Litz FW, Keane BT, Palmieri TM, Marx BP, Schnurr PP. The PTSD Checklist for DSM-5 (PCL-5). 2013.

[28] Connor KM, Davidson JRT, Churchill LE, Sherwood A, Weisler RH, Foa E. Psychometric properties of the Social Phobia Inventory (SPIN): New self-rating scale. Br J Psychiatry 2000;176:379–86. 10.1192/bjp.176.4.379.

[29] Turner GM, Litchfield I, Finnikin S, Aiyegbusi OL, Calvert M. General practitioners’ views on use of patient reported outcome measures in primary care: A cross-sectional survey and qualitative study. BMC Fam Pract 2020;21:14. 10.1186/s12875-019-1077-6.

[30] Connor KM, Kobak KA, Churchill LE, Katzelnick D, Davidson JRT. Mini-SPIN: A brief screening assessment for generalized social anxiety disorder. Depress Anxiety 2001;14:137–40. 10.1002/da.1055.

[31] Aminu AQ, Sinclair DR, Davies K, Hall A, Wiedemann M, O’Neill TW, et al. Disparities in the Use of Remote General Practice Consultations: Learning from the COVID-19 Pandemic, an Analysis of 19 Million Electronic Health Records Using OpenSAFELY 2025. 10.2139/ssrn.5174205.

[32] Davies B. Accurx Desktop - Questionnaire: SNOMED codes and questions for Depression PHQ-9 Questionnaire 2026. https://web.archive.org/web/20260914134110/ https://support.accurx.com/en/articles/768476-questionnaire-snomed-codes-and-questions-for-depression-phq-9-questionnaire (accessed September 18, 2026).

[33] Mundt JC, Marks IM, Shear MK, Greist JM. The Work and Social Adjustment Scale: A simple measure of impairment in functioning. Br J Psychiatry 2002;180:461–4. 10.1192/bjp.180.5.461.

[34] Delgadillo J, McMillan D, Gilbody S, de Jong K, Lucock M, Lutz W, et al. Cost-effectiveness of feedback-informed psychological treatment: Evidence from the IAPT-FIT trial. Behav Res Ther 2021;142:103873. 10.1016/j.brat.2021.103873.

[35] Wiedemann M, Saunders R, Thew G, Clark D, Ojedele L, Kingsley V, et al. nhstt: An openly accessible tool for analysing publicly available NHS Talking Therapies reports. Manuscr Prep 2026.

